# Workplace transmission of SARS-CoV-2 among health-care workers in Malaysia

**DOI:** 10.1101/2024.06.06.24306328

**Authors:** Chan Yean Yean, Muhammad Zarul Hanifah Md Zoqratt, Nyok-Sean Lau, Nurfadhlina Musa, Muhammad Fazli Khalid, Lim Shu Yong, Wan Mohd Zahiruddin Wan Mohammad, Sadequr Rahman, Wilhelm Wei Han Eng, Zaini bin Hussin, Abdul Haris bin Muhammad, Maizun Binti Mohd Zain, Suhaiza Binti Sulaiman, Nik Mohd Hafiz Bin Mohd Fuzi, Noor Hafizan Binti Mat Salleh, Syahida binti Omar, Ahmad Sukari Halim, Zakuan Zainy Deris, Kirnpal Kaur Banga Singh, Azian Harun, Muhammad Nashrul Farhan Samsudin, Chua Wei Lian, Nazmi Liana Binti Azmi, Engku Nur Syafirah bt Engku Abd Rahman, Naveed Ahmed, Nor Azwany Yaacob, Aliff Ridzwan Hamidun, Mohd Hazwan Baharuddin, Mohd Nasrullah Nik Ab. Kadir, Munira Hj Mahmud, Mohd Azmi Suliman, Qasim Ayub, Rosline Hassan

**Affiliations:** Department of Medical Microbiology and Parasitology, School of Medical Sciences, Universiti Sains Malaysia, Kubang Kerian 16150, Kelantan, Malaysia; Institute for Research in Molecular Medicine (INFORMM), Universiti Sains Malaysia, 16150 Kubang Kerian, Kelantan, Malaysia; Monash University Malaysia Genomics Facility, School of Science, Jalan Lagoon Selatan, 47500 Bandar Sunway, Selangor Darul Ehsan, Malaysia; Centre for Chemical Biology, Universiti Sains Malaysia, Bayan Lepas, 11900 Penang, Malaysia; Human Genome Centre, School of Medical Sciences, Universiti Sains Malaysia, 16150 Kubang Kerian, Kelantan, Malaysia; Department of Community Medicine, School of Medical Sciences, Universiti Sains Malaysia, 16150 Kubang Kerian, Kelantan, Malaysia; Kelantan State Health Department, Kelantan, Malaysia; Head of Public Health Unit HRPZ2, Medical Division, Kelantan State Health Department, Kelantan, Malaysia; Surveillance Unit, Kelantan State Health Department, Kelantan, Malaysia; Communicable Disease Control (CDC) Unit, Kelantan State Health Department, Malaysia; Kota Bharu Public Health Laboratory, Kelantan, Malaysia; Molecular lab, Disease Division, Kota Bharu Public Health Laboratory, Jalan Kuala Krai, 16010 Kota Bharu, Kelantan, Malaysia; Hospital Universiti Sains Malaysia, Universiti Sains Malaysia, Kubang Kerian 16150, Kelantan, Malaysia; Reconstructive Sciences Unit, School of Medical Sciences, Universiti Sains Malaysia, Kubang Kerian 16150, Kelantan, Malaysia; Department of Pharmacy, Raja Perempuan Zainab II Hospital, Ministry of Health Malaysia, Kota Bharu, Kelantan, Malaysia; Department of Haematology, School of Medical Sciences, Universiti Sains Malaysia, 16150, Kubang Kerian, Kelantan, Malaysia

**Keywords:** COVID-19, viral genomics, molecular epidemiology, genetic cluster

## Abstract

COVID19 genomic surveillance is instrumental to better understand transmission dynamics in a setting, detect emergence of new variants and monitor spread of variants at the national, regional and global levels. Complete viral genome sequences are powerful enough to approximate epidemiology and enable informed public health response policies and determine their success. Between 24th November to 9th December 2020, a workplace COVID-19 outbreak, assigned as the Hilir cluster, occurred among healthcare workers (HCWs) in a northeast Malaysian university teaching hospital that was not designated for COVID-19 treatment. Mass screening of 1,292 individuals based on case interviews, contact tracings and nucleic acid testing detected 17 cases from various hospital wards and units. To investigate how COVID19 transmission occurred we whole genome sequenced 14 samples collected from healthcare workers and 5 samples from the concurrent community outbreaks. The genomes of these samples were compared with closely-related publically available genomes from GISAID to gain insights into COVID19 transmission in the hospital and at the local and global scale. The 14 viral sequences obtained from the Hilir cluster were assigned to Pango lineages B.1.524 (7 samples) and B.1.36.16 (7 samples) whereas the community samples were assigned as either B.1.524 and B.1. Phylogenetics revealed multiple introduction of B.1.524 into the workplace, while close relatedness of all B.1.36.16 samples suggested that the introduction of these lineages into the workplace likely stemmed from a single introduction. These lines of genomic evidences contradicted with the proposed transmission route, underlining the central role of genomics in COVID-19 or any future pandemics surveillance. The study also highlight the difficulty in enforcing and maintaining isolation methods in a hospital setting.

**Impact statement:** Epidemiological outbreak investigation and sequencing of SARS-CoV-2 viral genomes was employed to study a healthcare workplace outbreak of COVID-19 that occurred in a Northeast Malaysian hospital between 24th November to 9th December 2020. We sought to understand the transmission dynamics of this Hilir cluster by sequencing samples obtained from 14 infected HCW and five infected individuals from the local community. The phylogeny of the viral sequences was studied using publically available SARS-CoV-2 genomes. We found two major COVID-19 lineages with possibly different transmission dynamics. The B.1.524 lineage was widespread in the community and possibly entered the workplace more than once whereas the B.1.36.16 lineages clustered together indicating a single introduction into the workplace. We also investigated COVID-19 transmission of the two lineages in the global context and found contradicting results between Pango lineage assignment and phylogenetic relatedness of B.1.36.16 genomes of Thailand-Malaysia-Singapore versus Bangladesh. This underlines the importance of integrating whole viral genome phylogenetics to complement epidemiological investigations and understand the spread of a disease in the community and guide national policies during pandemics.

**Outcome:**

⍰ The 14 sequences from the Hilir cluster and five community outbreak samples fall into two Pango lineages B.1.524 and B.1.36.16.
⍰ Phylogenetic analyses suggest that B.1.524 possibly entered the workplace multiple times as evidenced by distant phylogenetic relatedness of B.1.524 Hilir genomes into four separate subclades. The community samples were clustered within a B.1.524 subclade which also includes three samples from the Hilir healthcare workers cluster.
⍰ The viral sequence relatedness of B.1.36.16 Hilir genomes suggest a single entry into the workplace. None of the community samples were assigned as B.1.36.16

**Data summary:** Details on public genomes, metadata used and contributing authors are available from GISAID using EPI_SET ID EPI_SET_230904pv (Pango lineage B.1.524), EPI_SET_230904ps (Pango lineage B.1.36) and EPI_SET_230904ky (Pango lineage B.1.36.16).

The genome sequences generated from this projectare accessible through GISAID and GenBank accession IDs; refer to Supplementary Table 1. Full details and reproduction of the genome assembly and variant analysis is accessible through the Github repository: https://github.com/ZarulHanifah/snakemake_COVID19.

## Introduction

Coronavirus disease 2019 (COVID-19) was declared a Public Health Emergency of International Concern by the World Health Organization (WHO) on 30^th^ January 2020 [1, 2]. COVID-19 was first confirmed in Malaysia among travellers from China to Johor, in southern Peninsula Malaysia via Singapore on 25^th^ January 2020 [3]. The infection hit Malaysia in three waves. The first wave lasted from 25^th^ January to 16^th^ February 2020 (Supplementary Figure 1). The second from 27^th^ February and 30^th^ June 2020. Subsequently,the third wave began on 8^th^ September 2020, marked by a sudden four-fold increase in cases within two months. This surge was primarily attributed due significant outbreaks from the Benteng Lahad Datu and Tembok clusters located in the states of Sabah and Kedah, respectively (https://covid-19.moh.gov.my).

Healthcare workers (HCWs) are at high risk and vulnerable to COVID-19 infection and may play a significant role in hospital transmissions. For instance, hospital outbreaks were responsible for around half of the confirmed MERS-CoV infections, with HCWs accounting for about 40% of the cases [4, 5]. While the outbreak was initially limited to imported cases, several local clusters also emerged, including workplace transmission clusters among HCWs especially in major hospitals beginning in late September 2020. Towards the end of December 2020, there were 1,771 HCWs (1.2% of all HCWs) in Malaysia who contracted COVID-19 since the beginning of the pandemic. There was an increase in cases among HCWs during the third wave which led to the recording of nearly 80% relative to prior HCWs COVID-19 cases. One possible explanation was that it was due to the super-spreader characteristics of the SARS-CoV-2 virus identified in the third wave and the drastic rise in cases was believed to be closely linked with the increase in community transmissions. The HCWs were also suspected to acquire the infection not only through community transmissions but also from other HCWs and the transmission links were unclear [6].

This study aimed to understand a workplace transmission of COVID-19 at an epidemiologically-linked cluster in a tertiary care hospital in Northeast Malaysia that occurred in late 2020.

## Methods

### Ethical clearance

This study was approved by the Human Research Ethics Committee, Universiti Sains Malaysia (USM/JEPeM/COVID19-44) and National Medical Research Registration, Malaysia (NMRR-20-1645-55277 (IIR)).

### Study design and sites

An epidemiological outbreak investigation was conducted in a non-designated COVID-19 university teaching hospital in Northeast Peninsula Malaysia after an index case was reported in late November 2020 (Figure 1, individual K1). This was later identified a cluster of workplace transmission among HCWs and their family members, an outbreak that was later declared by the Ministry of Health Malaysia as the “Hilir” cluster, and named after it was identified among the population living a nearby locality. The outbreak investigation was conducted by the COVID-19 Public Health Unit managed by Hospital Universiti Sains Malaysia.

**Figure 1.**
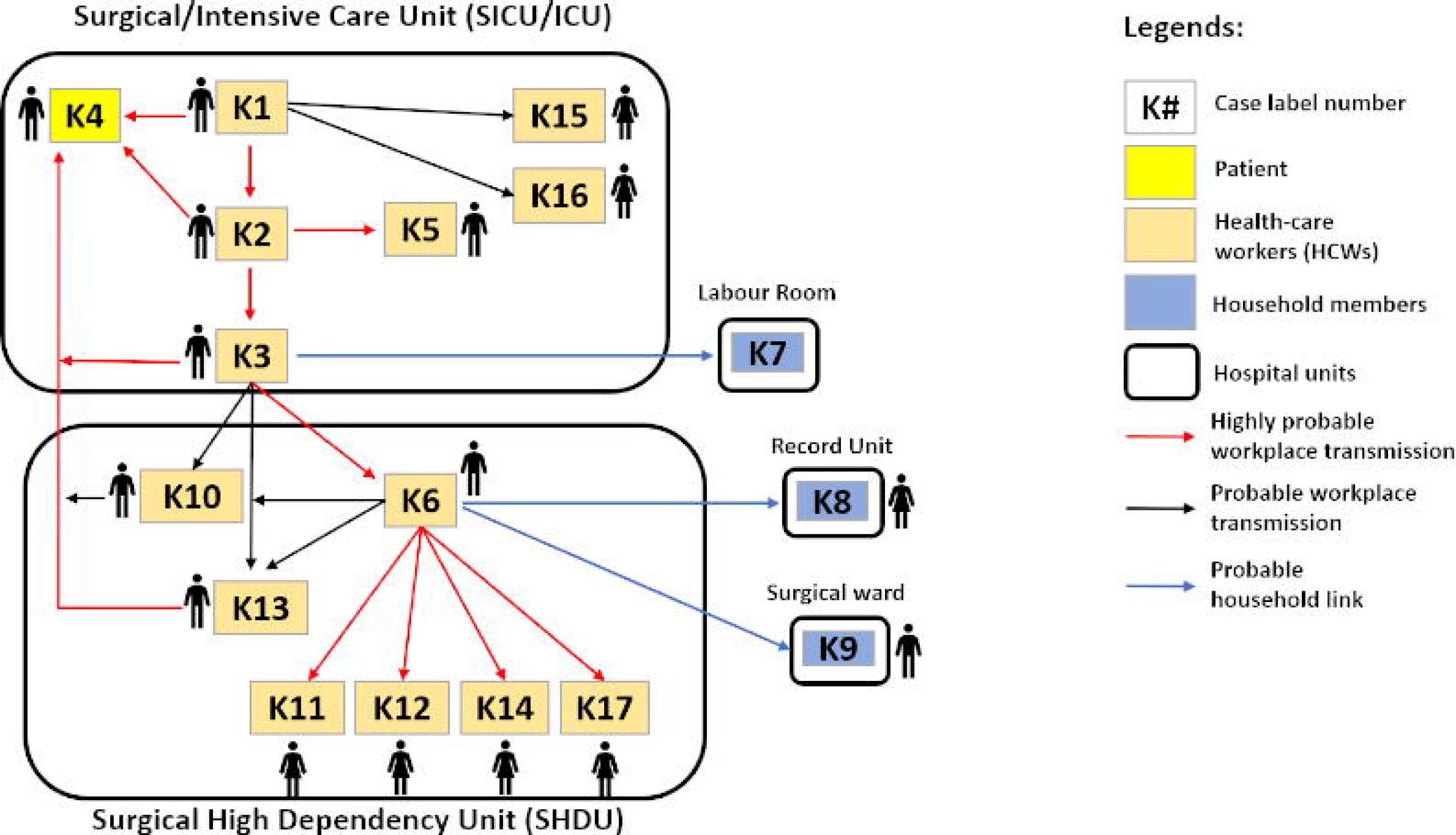
A) Probable epidemiological transmission among 17 COVID-19 Hilir cluster. B) Distribution of the 17 individuals from a Malaysia COVID-19 cluster investigated in this study. The HCWs were segregated according to their hospital departmental affiliations and date of diagnosis.

Prior to the outbreak, the hospital had no reported COVID-19 positive cases among HCWs as determined through its enhanced surveillance and screening of symptomatic workers and of those with recognized contact risk in the community. However, around the same time there were a few concurrent COVID-19 clusters within nearby communities, reported by the Ministry of Health. This includes local clusters called “Kube” declared on 7^th^ November 2020 with 17 cases;“Mengketil” identified on 17^th^ November 2020 with 24 cases, as well as the “Kaya” cluster detected on 24^th^ October 2020 with 55 cases (https://covid-19.moh.gov.my).

### Contact tracings and mass screenings

The index case (K1) of COVID-19 was determined using real-time reverse transcriptase-polymerase chain reaction (RT-PCR) from nasopharyngeal (NP) swabs. Close contacts of HCWs were defined as co-workers who worked in close proximity with the confirmed case during in-patient management, medical procedure trainings, or attendance in meetings held in closed areas or had social contact during work breaks, such as eating together in the pantry, or café, and sharing common rooms such as on-call and prayer rooms. Case interviews were done to determine the onset of symptoms. Asymptomatic cases were detected by close contact screening to identify high and medium risk close contacts based on the risk assessment guidelines. Such screening was conducted on close contact HCW as well as other non-staff household members or low risk contacts. Those who fulfilled the criteria of exposure included persons exposed to a case from 3 days prior to the onset of symptoms or, for asymptomatic cases, the day the swab was taken and screened using RT-PCR. High-risk contacts were home quarantined for 14 days from the last date of contact and repeat NP swabs were done after day 8, if the initial swab test result was negative for COVID-19 infection, As part of containment and surveillance strategies, mass screenings were conducted concurrently between 26^th^ November and 9^th^ December 2020 among all low risk HCWs from relevant hospital departments and units including symptomatic screenings. The department head and the nurse supervisors traced back the ward schedule and patients list to identify potential HCWs. Workplace activities, mapping as well as personal information were analysed to find out possible epidemiological link between the cases either within or outside the same workplaces. A total of 1,292 HCWs, including household members, were screened during this process.

All individuals consented to participate in the study. For those younger than 16 years permission of a parent or legal guardian was obtained

### Clinical presentation staging

The severity and five clinical stages, or categories of COVID-19 infection were determined based on the National Ministry of Health classification guidelines, Malaysia [7]; These included; Category 1, asymptomatic; Category 2, Symptomatic, No Pneumonia; Category 3, Symptomatic, Pneumonia; Category 4, Symptomatic, Pneumonia, Requiring supplemental oxygen; and Category 5, Critically ill with multi-organ involvement.

### Clinical specimens and RT-qPCR detection of SARS-CoV-2

The NP swab specimens were collected in viral transport medium (VTM) and screened using Lytestar 2019-nCoV RT-PCR Kit (IDT, Malaysia) which targets the viral *E* and *RdRP* genes with an internal control to rule out false negative results for confirmation. Fourteen SARS-CoV-2-positive Hilir cluster samples were positive and were later subjected to whole genome sequencing (WGS). In addition, five community samples from three local clusters (“Kube”, “Mengketil” and “Kaya”) detected at the same time point as the “Hilir” cluster (between November 2020 to December 2020) were also detected to be positive for SARS-CoV-2 and were included in the whole genome sequencing.

### SARS-CoV-2 genome sequencing

RNA was extracted from SARS-CoV-2 positive samples using the QIAamp viral RNA minikit (QIAGEN, Germany). The sequencing libraries were prepared following the ARTIC SARS-CoV-2 V3 amplification protocol (https://artic.network/ncov-2019) [8]. Briefly, cDNA was synthesized using the SuperScript IV first-strand synthesis system (Invitrogen, USA) with random hexamers and PCR was performed using Q5 Hot Start High-Fidelity DNA Polymerase (NEB, USA) with two pools of ARTIC V3 primers targeting the SARS-CoV-2 genome. The amplicons were combined, purified using AMPure XP beads (Beckman Coulter, USA), quantified with a Qubit 2.0 fluorometer and subjected to library construction using Nextera XT Library Prep Kit (Illumina, USA). Sequencing was performed on an Illumina MiSeq System (San Diego, CA, USA) with 2L×L250-bp paired-end reads configuration.

### Genome assembly and variant analysis

Raw sequencing data was trimmed to remove adapter sequences and low-quality bases using Trimmomatic (version 0.39) [9] and mapped to the reference genome Wuhan-Hu-1 (NC_045512.2) using Bowtie2 (version 2.4.2) [10]. The consensus genome sequences were generated from the aligned BAM files using iVar (version 1.3) [11]. The genomic sequences from this study were deposited in GISAID and GenBank (Supplementary Table 1). The consensus genomes were assigned to lineages with the PANGOLIN software (version 3.1.16) with pangolearn 2021-11-09 [12]. The BAM files were sorted by SAMtools (version 1.11) [13] and variants were called by BCFtools mpileup and call functions (version 1.11) [14], assuming a ploidy of 1. Full details and reproduction of the analysis is accessible through the Github repository: https://github.com/ZarulHanifah/snakemake_COVID19.

### Phylogenetic analysis

Phylogenetic tree for lineages B.1.524 and B.1.36.16 lineages were performed using all genomes of corresponding lineages from the GISAID database at date 15^th^ April 2022, using the Nextstrain SARS-CoV-2 workflow on Augur (version 14.0.0) [15]. Briefly, genome sequences were aligned to the reference genome using mafft (version 7.490) [16] against the Wuhan-Hu-1 reference genome. Then, phylogenetic trees were constructed using IQ-TREE (version 2.2.0_beta) [17]. A time tree was generated by adjusting the phylogenetic tree branch lengths to reflect the date of sample collection using TreeTime, with parameters substitution-model = GTR, clock_rate = 0.0008 per site per year, clock_std_dev = 0.0004, coalescent = opt, date_inference = marginal, divergence_unit = mutations, clock_filter_iqd = 8, keep-polytomies = True and no_covariance = True [18]. Annotations were further incorporated into the tree such as ancestral traits, nucleotide mutations and amino acid mutations. The final tree was then visualised using Auspice (version 2.29.1) [19] and Baltic (version 0.1.6).

## Results

### Epidemiological investigation finding

The index case (K1) was a HCW in their 50s whose onset of symptoms occurred in the middle of November 2020. The personnel underwent screening and was subsequently confirmed to be COVID-19 positive by RT-qPCR. Close contacts among his colleagues and clinical teams, meeting participants, selected patients and those sharing common places (pantries, on-call room and prayer room) within the hospital units were identified and screened for COVID-19. Between the end of November and early December 2020, a total of 17 HCWs were diagnosed to be infected with COVID-19 (Figure 1). Their sociodemographic characteristics are shown in Table 1. Epidemiological data of the confirmed cases revealed that majority of cluster cases were from the intensive care units (ICU) and surgical ICU (SICU) as well as the surgical high dependency unit (SHDU). The HCW categories ranged from specialist, staff nurses, medical assistants and ward medical attendants. Meanwhile, two HCWs (K7 and K9) were not working in either ICU, SICU or SHDU. Also, one non-HCW (K8), working in the record unit of the hospital is a household member of two HCWs (K3 and K6) who were found to be positive during the close contact screening. Figure 1 shows the probable epidemiological transmission among HCWs in various hospital units during the outbreak.

### Identification of B.1.524 and B.1.36.16 lineages in the Hilir cluster

SARS-CoV-2 genome sequencing results of the cluster “Hilir” individuals and concurrent community-based samples are shown in Supplementary Table 1. Three of the samples (K4, K15, and K17) were unavailable for sequencing analysis.

The PANGOLIN database grouped the Hilir cluster samples into two major lineages; the B.1.36.16 (7 samples) and the B.1.524 lineages (7 samples). The community samples Kube1, Mengketil2 and Kaya1 fells under the B.1524 lineage, whereas Mengketil1 and Kaya2 belongs to the B.1 lineage. No community samples were assigned as B.1.36.16. This agrees with the fact that between October and December of 2020, B.1.524 was the most prevalent lineage at 44.8 % in Malaysia, followed by B.1.36.16 at 10.4 % (Supplementary Figure 1). The high prevalence of the two lineages suggests their entry from the community into the workplace setting.

The detection of these two lineages suggests at least two introductions into the workplace had to happen, contradicting proposed epidemiological route in Figure 1. This also highlights the power of whole genome sequencing in confirming and complementing epidemiological approximation.

### Multiple entrance of B.1.524 into the workplace

The SAR-CoV-2 genome sequences generated from Kelantan, Malaysia were disproportionately low given the higher number of cases in the third wave as compared to the first two waves. Therefore, careful interpretation is required to determine whether the lineage distribution truly represents true Malaysian SARS-CoV-2 lineage distribution. Based on the genome sequences available, B.1.524 was the major lineage in Malaysia during the third wave (Supplementary Figure 1). Based on cov-lineages (https://cov-lineages.org), Malaysia has the highest number of genomes assigned as B.1.524 (74.8%), followed by neighbouring Singapore (12.9%), Thailand (4.2%) and Indonesia (1.0 %). This suggests that B.1.524 most likely originated from Malaysia and points towards the success of international travel ban to curb its spread into other countries (Figure 2).

**Figure 2.**
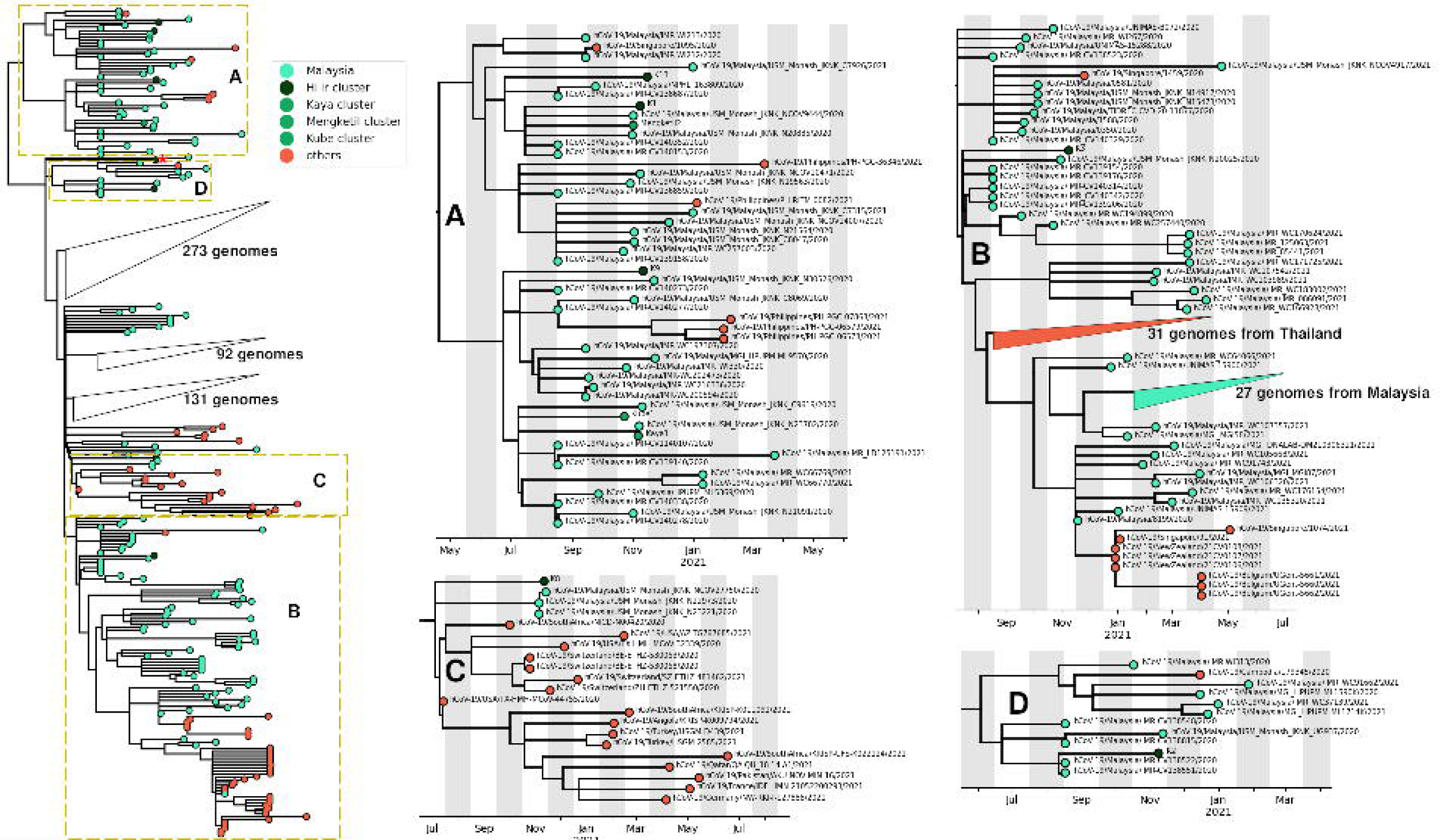
Phylogenetic tree of COVID19 genomes from GISAID with Pango lineage B.1.524. Many of genomes from the Hilir cluster are interspersed in the global B.1.524 tree. Different insets A-D display the different clades with genomes from the Hilir cluster. Sample K7 forms a singular polytomic branch, isolated from the other samples (red asterisk).

The B.1.524 lineage was first reported in USA Texas at 29^th^ July 2020 (Figure 2, clade C). Since clades A and D are basal to clade C, this suggests that the variant has been spreading around Malaysia even earlier (Figure 2). The B.1.524 lineage was first reported in Malaysia on 1^st^ September 2020, with as many as 22 genomes (Supplementary Figure 1). The basal root of the B.1.524 tree is predicted to originate at 9^th^ April 2020 (date confidence interval: 10^th^ August 2019 until 13^th^ June 2020) (Figure 2). The lack of genome sequencing efforts in Malaysia at the beginning of the pandemic could explains the failure to detect this lineage earlier, from April until September 2020.

Seven Hilir samples were assigned as B.1.524, but fall into four separate clades in the phylogenetic tree (Figure 2). Interstate spread cannot be inferred from samples generated by Institute for Medical Research (IMR) because the information was not made available in GISAID. There are other samples that were collected from the Kelantan state with sample IDs containing “USM_Monash_JKNK”, but was not part of the HCW surveillance effort. The results suggest at least four introductions of B.1.524 into workplace took place. If strictly each introduction forms a cluster that contains samples that are not distant by at most two weeks, then every B.1.524 Hilir sample comes from separate introductions. All Hilir samples have branch lengths of at least a bit over two months. These long isolated branches suggest undersampling, otherwise we would expect nodes that are closely related and sampled at around the same period. Clade A contains three Hilir samples and two community samples, all within November. There are also genomes from Singapore (October) and Phillippines (January until March 2021). Clade B contains sample K3. A single clade from Thailand was also found comprising 31 genomes. This suggests a singular introduction from Malaysia to Thailand, possibly before September. Later from January 2021, there is probably international spread of B.1.524 to other countries like Singapore, New Zealand and Belgium, spanning from January until May, although limited to only eight genomes. Clade C contains sample K8. This clade also contained genomes from other countries such as USA, South Africa, Switzerland, Turkey, Qatar, France, Pakistan and Germany. Finally Clade D contains sample K2.

### B.1.36.16 lineages in the Hilir cluster

B.1.36.16 was the second most prevalent SARS-CoV-2 lineage in the Malaysian third COVID-19 wave. Globally B.1.36.16 were reported from Bangladesh, Malaysia, Thailand and Singapore. Phylogenetic tree comprising of B.1.36 and B.1.36.16 genomes shows that the Bangladeshi samples are phylogenetically distant from those in Malaysia, Thailand and Singapore (Figure 3A). Focusing on the B.1.36.16 clade in the Southeast Asian region, we observed a strong regional clustering by country. We observed a broad distinct clustering of Malaysian clades and also a 98 genome clade exclusively from Thailand (Figure 3B). By scaling branches lengths to mutations, it is apparent that one of the Malaysian clades, including the samples from the Hilir cluster, accumulated 13 of mutations (Supplementary Figure 2). These nucleotide mutations include A1T, A4T, G11083T, G18756T, G27917A, G29881A, A29882G, G29884A, G29886A, A29887G, while the other three are reversion to reference genotype at positions 1947 (T), 25494 (T) and 29888 (G). Mutations X and Y resulted in two amino acid changes (X561V, L3606F) in ORF1a. This clade was sampled since August 2020 and includes 5 genomes that were obtained from Myanmar. Meanwhile, the 98-genomes Thailand clade was sampled from October 2020. Despite that this clade was found earlier, it accumulated more mutations than the Thailand clade. Narrowing down to clade containing Hilir samples, we found regional transmission patterns by state which are Kelantan and Negeri Sembilan (Figure 3C and 3D). Hilir samples fall under the Kelantan clade. This suggests success of interstate travel ban in curbing spread. Seven Hilir samples assigned as B.1.36.16 all fall into one single lineage with common ancestral node in November. The parent node to this clade dates back to August, indicating lack of genome-based surveillance especially in the third wave (Supplementary Figure 1). Otherwise, we expect to find genomes of this clade from August to November 2020.

**Figure 3.**
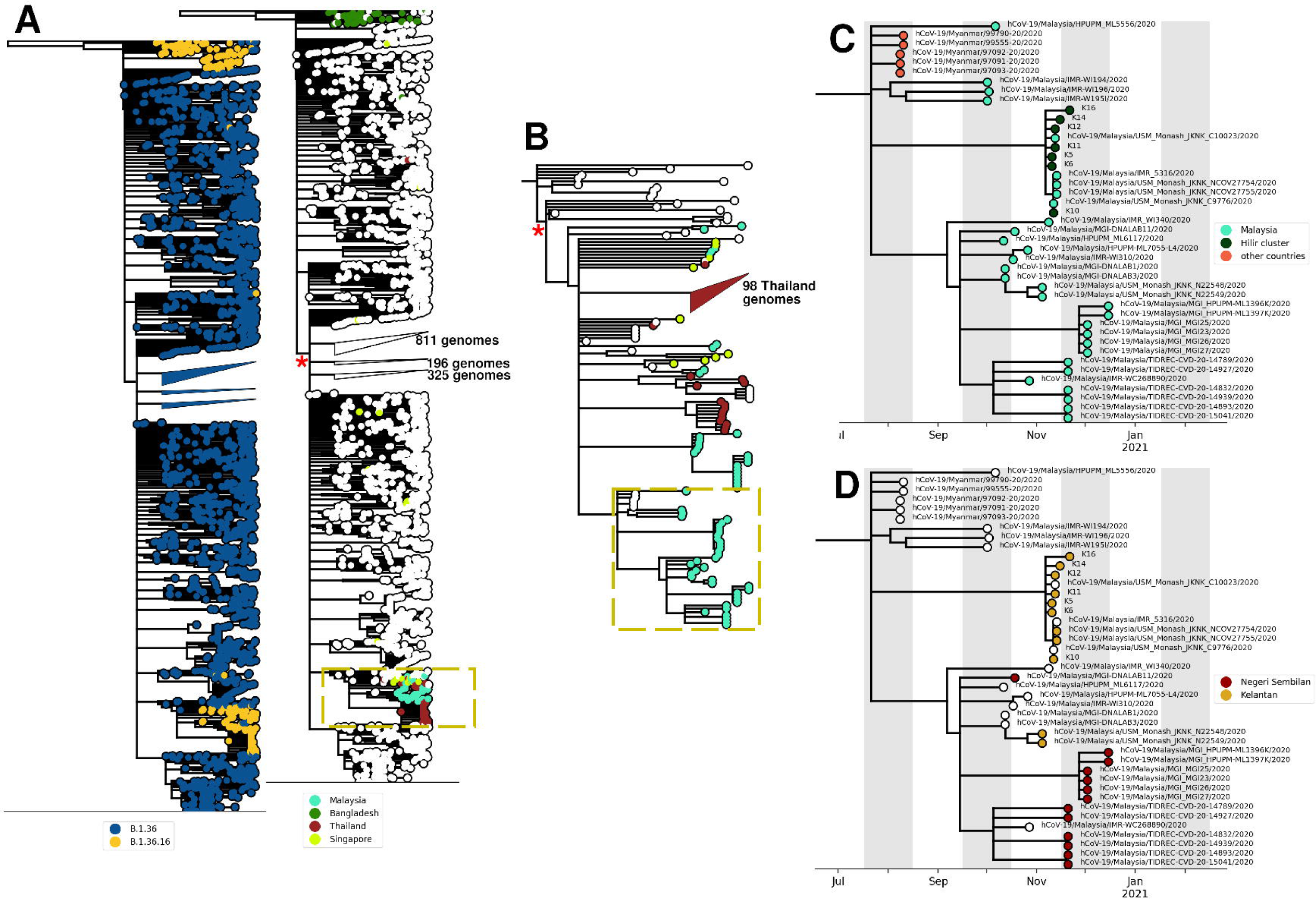
A) Phylogenetic tree of COVID19 genomes from GISAID with Pango lineage B.1.36 and B.1.36.16. The nodes of the left phylogenetic tree are coloured by Pango lineage, while the nodes of the right phlyogenetic tree are coloured by country of origin. B) Inset from (A), focusing on clade with B.1.36.16 Pango lineage, many from Malaysia, Thailand and Singapore. C) and D) refers to inset from Figure 3B. Nodes from Figure 3C phylogenetic tree are coloured by relation to this study, and also by country of origin.

### Variant analysis

Analysis of variants occurring in the SARS-CoV-2 strains sequenced in this study identified 147 non-synonymous mutations within the viral coding sequences, with most of the nucleotide substitutions located in the *ORF1ab* gene (Table 3). Mutations at amino acids P323L of the NSP12 protein, D614G of the spike (S) protein, and S194L of the nucleocapsid (N) protein were found to be most common in the strains sequenced in this study. Based on the phylogenetic tree, S194L arose independently in the two lineages B.1.524 and B.1.36.16 (https://genome.ucsc.edu/cgi-bin/hgPhyloPlace). Meanwhile, S194L of the N gene arose in B.1.36, which was later inherited to sub-lineages including B.1.36.16.

A 9-nucleotide sequence deletion was found at the position 656 to 664 (codon 131 of *nsp*1) in genomes K1, K2, K3, K9, K13, Mengketil2 and Kaya1 from the B.1.524 lineage, K6 and K11 from the B. 1.36.16 lineage, and Mengketil1 from the B.1 lineage.

### COVID-19 severity and spike protein gene mutations in the “Hilir” cluster

Based on the COVID-19’s severity, the majority (*n* = 8/14 or 57.1%) were categorised as Category 2 (symptomatic without pneumonia). Others were in Category 1 (*n*=3, 21.4%), Category 4 (*n*=1, 7.1%) and Category 5 (*n*=2, 14.3%). Both Category 5 COVID-19 workplace cluster HCWs were intubated, and one of them died after 12 days of hospital admission.

The epidemiological link and metadata such as clinical symptoms, date SARS-CoV-2 virus was detected, date of appearance of first symptoms, RT-PCR results, comorbidity background are provided by timeline and transmission chain in Table 4. The spike protein gene mutations of SARS-CoV-2 cluster “Hilir” clinical samples found the D614G mutation in all samples (100% of the 14 sequenced samples) and 79% of these samples had the A701V mutation [20].

## Discussion

COVID-19 workplace transmissions, especially among HCWs, mainly occurs through direct or indirect exposures to infected patients or their co-workers [21]. In view of concurrent COVID-19 transmission in the community, workplace transmission induced by community-infected HCWs is a major concern especially for non-COVID-19 designated hospitals [22, 23]. In this study, we conducted a thorough epidemiological outbreak investigation and management of a local cluster that involved prompt contact tracing, screenings and isolation of HCWs as well as enhanced surveillance and mass screenings.

Molecular diagnostic and viral genome sequencing approaches in combination with epidemiological data have proven to be instrumental for investigating outbreaks of COVID-19 in health-care settings [23, 24]. However, in Malaysia, viral sequencing has not been routinely used for investigating the cause of transmission and spread of this disease. The current study is among the first in the country. The focus of this study was on a HCWs cluster, termed “Hilir” cluster, that was identified in a state hospital in north eastern Malaysia. The cluster comprised of 17 individuals and complete viral sequences were obtained from 14/17 samples. Three of the samples were not sequenced because the sample was either insufficient (K4) or unavailable (K15 and K17).

The Hilir HCW cluster samples belonged to two major lineages, B.1.524 and B.1.36.16 and B.1.524. Both lineages were present in Malaysia between October-December 2020, with B.1.524 being the dominant lineage [25]. The presence of these two lineages suggests at least two introductions into the hospital and contradicts the proposed epidemiological route shown in Figure 1A.

The Hilir B.1.524 lineages fall into 4 separate clades (Figure 2) suggesting multiple introduction of the virus into the workplace and failure of the strict control measures enforced to stop the spread of COVID-19, The seven Hilir B.1.36.16 all share a common ancestral node indicating clonal relatedness between these cases. This highlights the power of whole genome sequencing in confirming and complementing epidemiological approximation.

Preliminary genomic analysis during the third wave in Malaysia showed the presence of the spike protein mutant variants D614G and A701V. These are typical of known super-spreader strains which caused an increase in the number of cases from 2,234 cases on 10^th^ December 2020 to 5,728 cases on 30^th^ January 2021. Since national level vaccination started on 1^st^ March 2021, the numbers have decreased to around 1200 cases per day as of April, 2021 [26]. A previous study conducted in the Netherlands used WGS for hospital outbreak investigations and demonstrated that hypothesis about viral transmission routes based exclusively on epidemiological data can be erroneous [27]. The same study also reported that infection control strategies can be improved with viral genome sequencing, especially if the results can be provided rapidly and compared to viral reference sequences currently available in different databases.

This study also found a clustering of cases from the ICU and high dependency unit where the first case mainly worked. HCWs at ICU and high dependency units had to comply with the SOPs on personal protective equipment (PPE) when handling patients. However, their compliance to workplace SOP during COVID-19 with other co-workers such as wearing face mask, physical distancing may not have been strict, especially during non-clinical duties and social contacts in common places such as pantries and prayer rooms. This increased the likelihood of community infection and most likely led to occupational workplace transmissions. Similar findings had been reported in a multinational study involving six Asian countries that earlier local transmission had resulted in a significant increase in occupational infections [28]. Several studies have also shown that around 85% of HCWs were exposed while wearing a usual surgical mask during an aerosol-generating procedure [27, 29] and current recommendations include wearing precise surgical masks (e.g., N95), hand hygiene, and other standard precautions to keep HCWs safe from this viral infection [30].

The probable epidemiological linkages of the cluster as shown in Figure 2 could improve the understanding of transmissibility of the virus based on workplace setting and exposure risk activities. However, the secondary attack rate could not be determined based on the incubation and infectivity period as well as the epidemic curve due to the concurrent mass screenings done among different levels of contacts, exposure risks and symptoms. The mass screening was able to identify cases among low-risk contacts, irrespective of whether they had evident occupational exposure to patients or co-workers, or possible linkages to concurrent community-based clusters. A similar study from the United States of America reported that the major risk factors associated with COVID-19 infection among HCWs are community attack rates and ethnicities [31].

NSP1 has been considered a key pathogenic determinant of coronaviruses that serves to suppress both host’s anti-viral innate immune response and global gene expression level [32]. We observed a 9 bp deletion in *nsp1* in 10/19 samples and its effect on viral function merit further investigation. The frequency of mutations within SARS-CoV-2 examined in this work could broadly classify these strains into two main groups: the B.1.524 strains were distinguished by the absence of Q57H mutation, and the B.1.36.16 strains by the absence of the T11981, T281, T24A and L428F mutations. K8 shows a distinct variants profile from the other B.1.524 strains by the absence of deletion in *nsp1* and T281 nonsynonymous mutation.

During the WHO and China Joint research mission on COVID-19, 2,055 laboratory confirmed cases were reported among HCWs from 476 hospitals across China, with Hubei province accounting for 88% of the cases. However, there was insufficient evidence for nosocomial infections and most of the HCWs were considered to have been infected in their homes rather than in the hospital [28]. The transmission of COVID-19 among HCWs does not necessarily indicate the non-compliance of SOPs inside the wards, operation theatres or procedure rooms. One of the reasons could be that there is comparatively less compliance of SOPs while the HCWs are outside their duty stations for example, during the lunch or tea breaks, and in the prayer rooms. The other risk of transmission could be when the HCWs visit outside places like shopping malls or picnic areas. To minimise workplace transmission, compliance with SOPs should be strictly maintained throughout the hospital settings. The HCWs should also minimise their social gatherings to reduce the chances of hospital transmissions.

We acknowledge the untimely release of this work, which is almost two years since the Hilir cluster. To enable real-time informed decision making, national effort to streamline COVID-19 analysis, dissemination of information and concerted research efforts are paramount.

### Conclusion

The Hilir cluster actually comprised of two SARS-CoV-2 lineages which are B.1.524 and B.1.36.16. Genomics-based pandemic surveillance plays an important role as a specific tool in understanding SARS-CoV-2 transmission, particularly the possibility of more than two transmission events of SARS-CoV-2 among a HCWs workplace cluster in Malaysia.

## Author statements Authors and contributors

**Conceptualization:** (C.Y.Y), (L.N.S), (M.Z.H.M.Z), (A.S.H), (A.H), (A.C.S.C), (Q.A.), (R.H.)

**Data curation:** (C.Y.Y), (M.Z.H.M.Z), (L.N.S), (N.M), (L.S.Y), (W.M.Z.W.M**), (**S.R), (W.E.W.H), (A.H), (C.W.L), (N.L.B.A), (A.C.S.C), (Q.A.), (R.H.)

**Formal analysis:** (C.Y.Y), (M.Z.H.M.Z), (L.N.S), (N.M), (M.F.B.K), (W.M.Z.W.M**), (**A.H), (N.A.Y), (A.R.H), (M.H.B), (M.N.N.A.K), (M.H.M), (A.M.S), (A.C.S.C), (Q.A.), (R.H.)

**Funding acquisition:** (C.Y.Y), (A.S.H), (Z.Z.D), (A.H), (A.C.S.C), (Q.A.), (R.H.)

**Investigation:** (C.Y.Y), (M.Z.H.M.Z), (L.N.S), (N.M), (M.F.B.K), (L.S.Y), (W.M.Z.W.M), (Z.B.H),, (A.H.B.M), (M.B.M.Z), (S.B.S), (N.M.H.B.M.F), (N.H.B.M.S), (S.B.O), (A.H), (M.N.F.S), (E.N.S.B.E.A.R), (N.A.Y), (A.R.H), (M.H.B), (M.N.N.A.K), (M.H.M), (A.M.S)

**Methodology:** (M.Z.H.M.Z, (L.N.S), (N.M), (M.F.B.K), (L.S.Y), (S.R), (W.E.W.H), (Z.B.H),, (A.H.B.M), (M.B.M.Z), (S.B.S), (N.M.H.B.M.F), (N.H.B.M.S), (S.B.O), (A.S.H), (Z.Z.D), (K.K.B.S), (A.H), (M.N.F.S), (C.W.L), (N.L.B.A), (E.N.S.B.E.A.R), (N.A.Y), (A.R.H), (M.H.B), (M.N.N.A.K), (M.H.M), (A.M.S)

**Project administration:** (C.Y.Y), (E.N.S.B.E.A.R), (A.C.S.C), (Q.A.), (R.H.)

**Resources:** (C.Y.Y), (N.M), (M.F.B.K), (W.M.Z.W.M), (Z.B.H),, (A.H.B.M), (M.B.M.Z), (S.B.S), (N.M.H.B.M.F), (N.H.B.M.S), (S.B.O), (A.S.H), (Z.Z.D), (K.K.B.S), (M.N.F.S), (C.W.L), (N.L.B.A), (A.C.S.C), (Q.A.), (R.H.)

**Software:** (M.Z.H.M.Z), (L.N.S)

**Supervision:** (A.C.S.C), (Q.A.), (R.H.)

**Validation:** (C.Y.Y), (M.Z.H.M.Z), (L.N.S), (N.M), (L.S.Y), (S.R), (W.E.W.H), (A.H), (A.C.S.C), (Q.A.), (R.H.)

**Visualization:** (C.Y.Y), (N.M), (A.H), (N.A)

**Writing – original draft:** (C.Y.Y), (L.N.S), (N.M), (W.M.Z.W.M**), (**A.H), (N.A), (N.A.Y), (R.H.)

**and Writing – review & editing:** (C.Y.Y), (M.Z.H.M.Z), (W.M.Z.W.M**), (**S.R), (W.E.W.H), (A.S.H), (K.K.B.S), (A.H), (N.A), (N.A.Y), (A.C.S.C), (Q.A.), (R.H.)

## Ethical approval

This study was approved by the Human Research Ethics Committee, Universiti Sains Malaysia (USM/JEPeM/COVID19-44) and National Medical Research Registration, Malaysia (NMRR-20-1645-55277 (IIR)).

## Data Availability

All data produced are available online at GISAID using EPI_SET ID EPI_SET_230904pv (Pango lineage B.1.524), EPI_SET_230904ps (Pango lineage B.1.36) and EPI_SET_230904ky (Pango lineage B.1.36.16)

https://gisaid.org/

## Acknowledgements

All authors would like to acknowledge the HUSM for the SARS-COV-2 laboratory facilities. M.Z.H.M.Z was supported by Monash Malaysia R&D Sdn. Bhd. E.N.S.B.E.A.R was supported by the USM Fellowship Scheme. N.A was supported by USM’s Graduate Research Assistant Scheme.

---

**Supplementary Figure 1.**
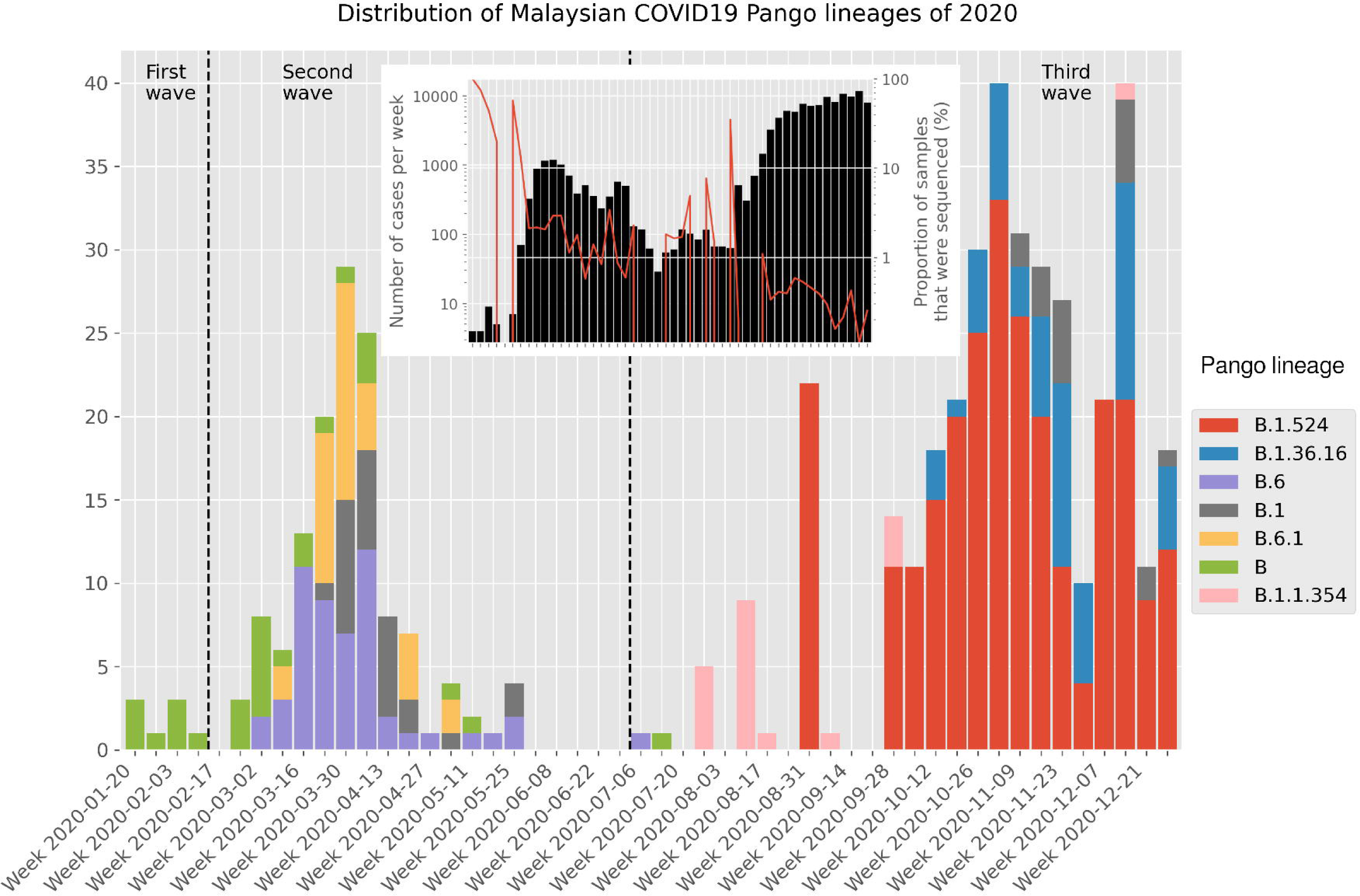
Distribution of major Malaysian COVID19 Pango lineages in 2020, obtained from GISAID. Inset: Barplots displaying the number of new cases per week in Malaysia, obtained from Our World in Data (https://ourworldindata.org/). The red line shows the proportion of samples that were sequenced at that week. Both y-axis are log-transformed.

**Supplementary Figure 2.**
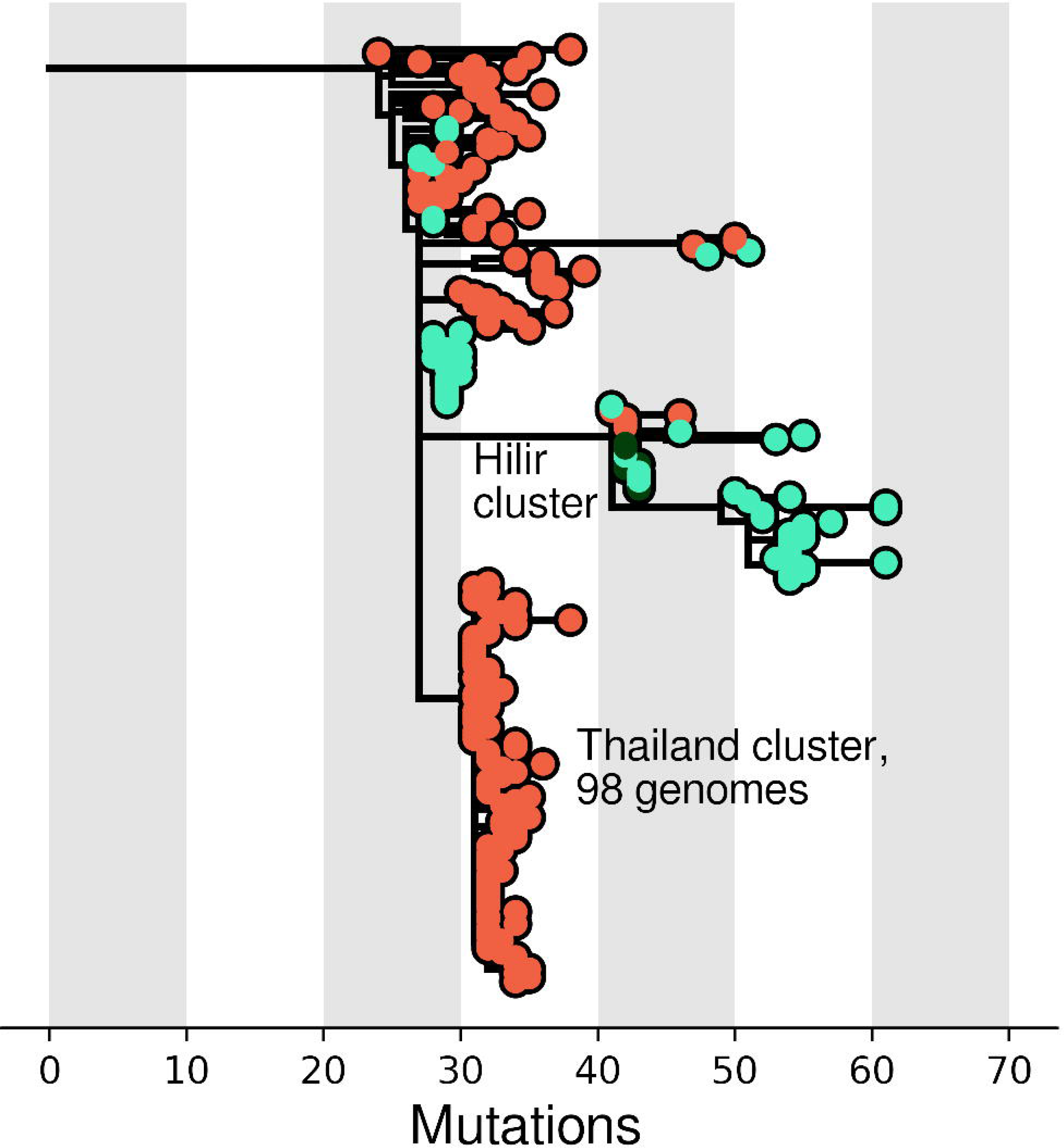
Inset from Figure 3B, x-axis represents number of mutations

**Supplementary Table 1.** SARS-CoV-2 genome sequencing of cluster “Hilir” and concurrent community-based clusters. Asterisks refer to cases of concurrent community-based cluster.

**Supplementary Table 2.** Mutation profiles of 19 Malaysian SARS-CoV-2 genomes obtained from the Hilir (n = 14) and community (n = 5) clusters. Samples were sorted by Pango lineage. Asterisks refer to non-Hilir cluster.

## References

1. Koh D, Goh HP. Occupational health responses to COVID-19: What lessons can we learn from SARS? Journal of occupational health 2020;62:e12128.

2. Musa KI, Arifin WN, Mohd MH, Jamiluddin MS, Ahmad NA, et al. Measuring Time-Varying Effective Reproduction Numbers for COVID-19 and Their Relationship with Movement Control Order in Malaysia. Int J Environ Res Public Health;18. Epub ahead of print March 2021. DOI: 10.3390/ijerph18063273.

3. Khoo LS, Hasmi AH, Ibrahim MA, Mahmood MS. Management of the dead during COVID-19 outbreak in Malaysia. Forensic Sci Med Pathol 2020;16:463–470.

4. Niemeyer D, Mösbauer K, Klein EM, Sieberg A, Mettelman RC, et al. The papain-like protease determines a virulence trait that varies among members of the SARS-coronavirus species. PLoS Pathog 2018;14:e1007296.

5. Rivett L, Sridhar S, Sparkes D, Routledge M, Jones NK, et al. Screening of healthcare workers for SARS-CoV-2 highlights the role of asymptomatic carriage in COVID-19 transmission. Elife;9. Epub ahead of print May 2020. DOI: 10.7554/eLife.58728.

6. Safuan S, Edinur HA. Sri Petaling COVID-19 cluster in Malaysia: challenges and the mitigation strategies. Acta bio-medical: Atenei Parmensis 2020;91:e2020154.

7. Rampal L, Liew BS. Malaysia’s third COVID-19 wave - a paradigm shift required. The Medical journal of Malaysia 2021;76:1–4.

8. Tyson JR, James P, Stoddart D, Sparks N, Wickenhagen A, et al. Improvements to the ARTIC multiplex PCR method for SARS-CoV-2 genome sequencing using nanopore. Epub ahead of print September 2020. DOI: 10.1101/2020.09.04.283077.

9. Bolger AM, Lohse M, Usadel B. Trimmomatic: a flexible trimmer for Illumina sequence data. Bioinformatics 2014;30:2114–2120.

10. Langmead B, Salzberg SL. Fast gapped-read alignment with Bowtie 2. Nat Methods 2012;9:357–359.

11. Grubaugh ND, Gangavarapu K, Quick J, Matteson NL, Jesus JG De, et al. An amplicon-based sequencing framework for accurately measuring intrahost virus diversity using PrimalSeq and iVar. Genome Biol;20. Epub ahead of print 2019. DOI: 10.1186/s13059-018-1618-7.

12. O’Toole Á, Scher E, Underwood A, Jackson B, Hill V, et al. Assignment of epidemiological lineages in an emerging pandemic using the pangolin tool. Virus Evol;7. Epub ahead of print 2021. DOI: 10.1093/ve/veab064.

13. Li H, Handsaker B, Wysoker A, Fennell T, Ruan J, et al. The Sequence Alignment/Map format and SAMtools. Bioinformatics 2009;25:2078–2079.

14. Danecek P, Bonfield JK, Liddle J, Marshall J, Ohan V, et al. Twelve years of SAMtools and BCFtools. Gigascience;10. Epub ahead of print 2021. DOI: 10.1093/gigascience/giab008.

15. Huddleston J, Hadfield J, Sibley TR, Lee J, Fay K, et al. Augur: a bioinformatics toolkit for phylogenetic analyses of human pathogens. J Open Source Softw 2021;6:2906.

16. Katoh K, Standley DM. MAFFT multiple sequence alignment software version 7: Improvements in performance and usability. Mol Biol Evol. Epub ahead of print 2013. DOI: 10.1093/molbev/mst010.

17. Minh BQ, Schmidt HA, Chernomor O, Schrempf D, Woodhams MD, et al. IQ-TREE 2: New Models and Efficient Methods for Phylogenetic Inference in the Genomic Era. Mol Biol Evol 2020;37:1530–1534.

18. Sagulenko P, Puller V, Neher RA. TreeTime: Maximum-likelihood phylodynamic analysis. Virus Evol 2018;4:vex042.

19. Hadfield J, Megill C, Bell SM, Huddleston J, Potter B, et al. Nextstrain: real-time tracking of pathogen evolution. Bioinformatics 2018;34:4121–4123.

20. Korber B, Fischer WM, Gnanakaran S, Yoon H, Theiler J, et al. Tracking Changes in SARS-CoV-2 Spike: Evidence that D614G Increases Infectivity of the COVID-19 Virus. Cell 2020;182:812–827.e19.

21. Black JRM, Bailey C, Przewrocka J, Dijkstra KK, Swanton C. COVID-19: the case for health-care worker screening to prevent hospital transmission. Lancet (London, England) 2020;395:1418–1420.

22. Lan F-Y, Wei C-F, Hsu Y-T, Christiani DC, Kales SN. Work-related Covid-19 transmission. medRxiv. Epub ahead of print 2020. DOI: 10.1101/2020.04.08.20058297.

23. Heinzerling A, Stuckey MJ, Scheuer T, Xu K, Perkins KM, et al. Transmission of COVID-19 to Health Care Personnel During Exposures to a Hospitalized Patient Solano County, California, February 2020. Morb Mortal Wkly Rep 2020;69:472–476.

24. Treibel TA, Manisty C, Burton M, McKnight Á, Lambourne J, et al. COVID-19: PCR screening of asymptomatic health-care workers at London hospital. Lancet (London, England) 2020;395:1608–1610.

25. Zainulabid UA, Mat Yassim AS, Hussain M, Aslam A, Soffian SN, et al. Whole genome sequence analysis showing unique SARS-CoV-2 lineages of B.1.524 and AU.2 in Malaysia. PLoS One 2022;17:e0263678.

26. Aw SB, Teh BT, Ling GHT, Leng PC, Chan WH, et al. The COVID-19 Pandemic Situation in Malaysia: Lessons Learned from the Perspective of Population Density. International journal of environmental research and public health;18. Epub ahead of print June 2021. DOI: 10.3390/ijerph18126566.

27. Sikkema RS, Pas SD, Nieuwenhuijse DF, O’Toole Á, Verweij J, et al. COVID-19 in health-care workers in three hospitals in the south of the Netherlands: a cross-sectional study. Lancet Infect Dis 2020;20:1273–1280.

28. Lan F-Y, Wei C-F, Hsu Y-T, Christiani DC, Kales SN. Work-related COVID-19 transmission in six Asian countries/areas: A follow-up study. PLoS One 2020;15:e0233588.

29. Adams JG, Walls RM. Supporting the Health Care Workforce During the COVID-19 Global Epidemic. JAMA 2020;323:1439–1440.

30. Hurst KR, Koetzner CA, Masters PS. Characterization of a critical interaction between the coronavirus nucleocapsid protein and nonstructural protein 3 of the viral replicase-transcriptase complex. J Virol 2013;87:9159–9172.

31. Lan F-Y, Filler R, Mathew S, Buley J, Iliaki E, et al. Sociodemographic risk factors for coronavirus disease 2019 (COVID-19) infection among Massachusetts healthcare workers: A retrospective cohort study. Infect Control Hosp Epidemiol 2021;42:1473– 1478.

32. Yuan S, Balaji S, Lomakin IB, Xiong Y. Coronavirus Nsp1: Immune Response Suppression and Protein Expression Inhibition. Front Microbiol 2021;12:752214.

